# Mapping the relationship of white matter lesions to depression in multiple sclerosis

**DOI:** 10.1101/2023.06.09.23291080

**Authors:** Erica B. Baller, Elizabeth M. Sweeney, Matthew C. Cieslak, Timothy Robert-Fitzgerald, Sydney C. Covitz, Melissa L. Martin, Matthew K. Schindler, Amit Bar-Or, Ameena Elahi, Bart S. Larsen, Abigail R. Manning, Clyde E. Markowitz, Christopher M. Perrone, Victoria Rautman, Madeleine M. Seitz, John A. Detre, Michael D. Fox, Russell T. Shinohara, Theodore D. Satterthwaite

**Author notes:** **Corresponding author:** Theodore D. Satterthwaite, M.D., Richards Building, 5th Floor, Suite 5A, 3700 Hamilton Walk, Philadelphia, PA 19104-6085. co-senior authors who contributed equally (Russell T. Shinohara, Theodore D. Satterthwaite).

## Abstract

**Importance:** Multiple sclerosis (MS) is an immune-mediated neurological disorder that affects nearly one million people in the United States. Up to 50% of patients with MS experience depression.

**Objective:** To investigate how white matter network disruption is related to depression in MS.

**Design:** Retrospective case-control study of participants who received research-quality 3-tesla neuroimaging as part of MS clinical care from 2010-2018. Analyses were performed from May 1 to September 30, 2022.

**Setting:** Single-center academic medical specialty MS clinic.

**Participants:** Participants with MS were identified via the electronic health record (EHR). All participants were diagnosed by an MS specialist and completed research-quality MRI at 3T. After excluding participants with poor image quality, 783 were included. Inclusion in the depression group (*MS+Depression*) required either: 1) ICD-10 depression diagnosis (F32-F34.*); 2) prescription of antidepressant medication; or 3) screening positive via Patient Health Questionnaire-2 (PHQ-2) or -9 (PHQ-9). Age- and sex-matched nondepressed comparators (*MS-Depression*) included persons with no depression diagnosis, no psychiatric medications, and were asymptomatic on PHQ-2/9.

**Exposure:** Depression diagnosis.

**Main Outcome(s) and Measure(s):** We first evaluated if lesions were preferentially located within the depression network compared to other brain regions. Next, we examined if MS+Depression patients had greater lesion burden, and if this was driven by lesions specifically in the depression network. Outcome measures were the burden of lesions (e.g., impacted fascicles) within a network and across the brain. Secondary measures included between-diagnosis lesion burden, stratified by brain network. Linear mixed-effects models were employed.

**Results:** Three hundred-eighty participants met inclusion criteria, (232 MS+Depression: age[SD]=49[12], %females=86; 148 MS-Depression: age[SD]=47[13], %females=79). MS lesions preferentially affected fascicles within versus outside the depression network (β=0.09, 95% CI=0.08-0.10, P<0.001). MS+Depression had more white matter lesion burden (β=0.06, 95% CI=0.01-0.10, P=0.015); this was driven by lesions within the depression network (β=0.02, 95% CI 0.003-0.040, P=0.020).

**Conclusions and Relevance:** We provide new evidence supporting a relationship between white matter lesions and depression in MS. MS lesions disproportionately impacted fascicles in the depression network. MS+Depression had more disease than MS-Depression, which was driven by disease within the depression network. Future studies relating lesion location to personalized depression interventions are warranted.

**KEY POINTS:** *Question:* Are white matter lesions that impact fascicles connecting a previously-described depression network associated with depression in patients with multiple sclerosis (MS)?

*Findings:* In this retrospective, case-control study of patients with MS including 232 patients with depression and 148 nondepressed MS comparators, patients with MS had more disease within the depression network, irrespective of depression diagnosis. Patients with depression had more disease than those without depression, which was driven by disease within the depression network specifically.

*Meaning:* Lesion location and burden may contribute to depression comorbidity in MS.

## INTRODUCTION

Multiple sclerosis (MS) is an immune-mediated neurological disorder that affects nearly one million people in the United States^1,2^. It is characterized by demyelinating white matter lesions in the central nervous system^3–6^. Depression is highly comorbid with MS; up to 50% of patients with MS experience depression^7,8^. Depression in MS contributes to increased morbidity and mortality and is associated with suicide rates double that of persons without MS^9,10^. The rates of depression in MS are also higher than depression comorbidity in other chronic autoimmune diseases, suggesting that the neural pathophysiology of MS may confer increased depression risk^11^. Despite the overlap between MS and depression, their association is not well-understood^12^. The purpose of this study was to evaluate whether lesions affecting white matter tracts that connect brain regions previously associated with depression contribute to depression in MS.

Previous studies of medically healthy participants with depression have described associations between white matter properties and depressive symptoms in numerous cortical and subcortical white matter fascicles, though the directionality and strength of associations have been inconsistent^13,14^. However, research frequently excludes participants with intracranial pathology; thus, results from these studies cannot be extrapolated to MS. Small studies in MS that aimed to identify damage to fascicles associated with depression have also yielded mixed results^7, 15–17;^ injury to arcuate fasciculus, temporal, and superior and inferior frontal regions have been reported. Such heterogeneity may reflect study designs that evaluated fascicles individually rather than assessing how lesions in different anatomical locations within a functional network contribute to depressive symptoms. Techniques that explore how depression is associated with heterogeneous white matter disease in connected brain networks are vital to fill in this knowledge gap.

One approach that has proven powerful in relating heterogeneous lesions to the emergence of neuropsychiatric symptoms is *lesion network mapping* (LNM). LNM leverages normative human connectome data to identify functional circuits connected to a given lesion and compares circuits between patients with and without the diagnosis of interest (e.g., depression)^18,19^. In recent work, researchers demonstrated that strokes associated with depression are connected to a specific brain network^20,21^. Brain stimulation at sites within this “depression network” also alleviated depression symptoms. Finally, white matter lesions connected to this network have been linked to depression in MS using functional connectivity^22^. However, prior work has not directly characterized how injury to white matter fascicles may be linked to depression in MS. While white matter injury has been associated with overall disability in MS^23–25^, few studies have tested whether white matter lesion location and burden are associated with depression symptoms specifically.

In this study, we assessed the relationship between depression and white matter lesion location and burden in a large sample of patients with MS. Given the high degree of comorbidity between MS and depression, we hypothesized that MS lesions would preferentially target fascicles that connect the depression network. Furthermore, we predicted that MS patients with depression would have a greater lesion burden within the depression network than MS patients without depression.

## METHODS

### Participants

Participants with MS were identified from the electronic health record (EHR) via the University of Pennsylvania’s Data Analytic Center, including demographics, International Classification of Diseases (ICD)-10 diagnoses^26^, medication lists, and depression screens including Patient Health Questionnaires (two-question form, PHQ-2; and nine-question form, PHQ-9; **Figure 1**)^27^. The University of Pennsylvania Institutional Review Board approved this study.

**Figure 1.**
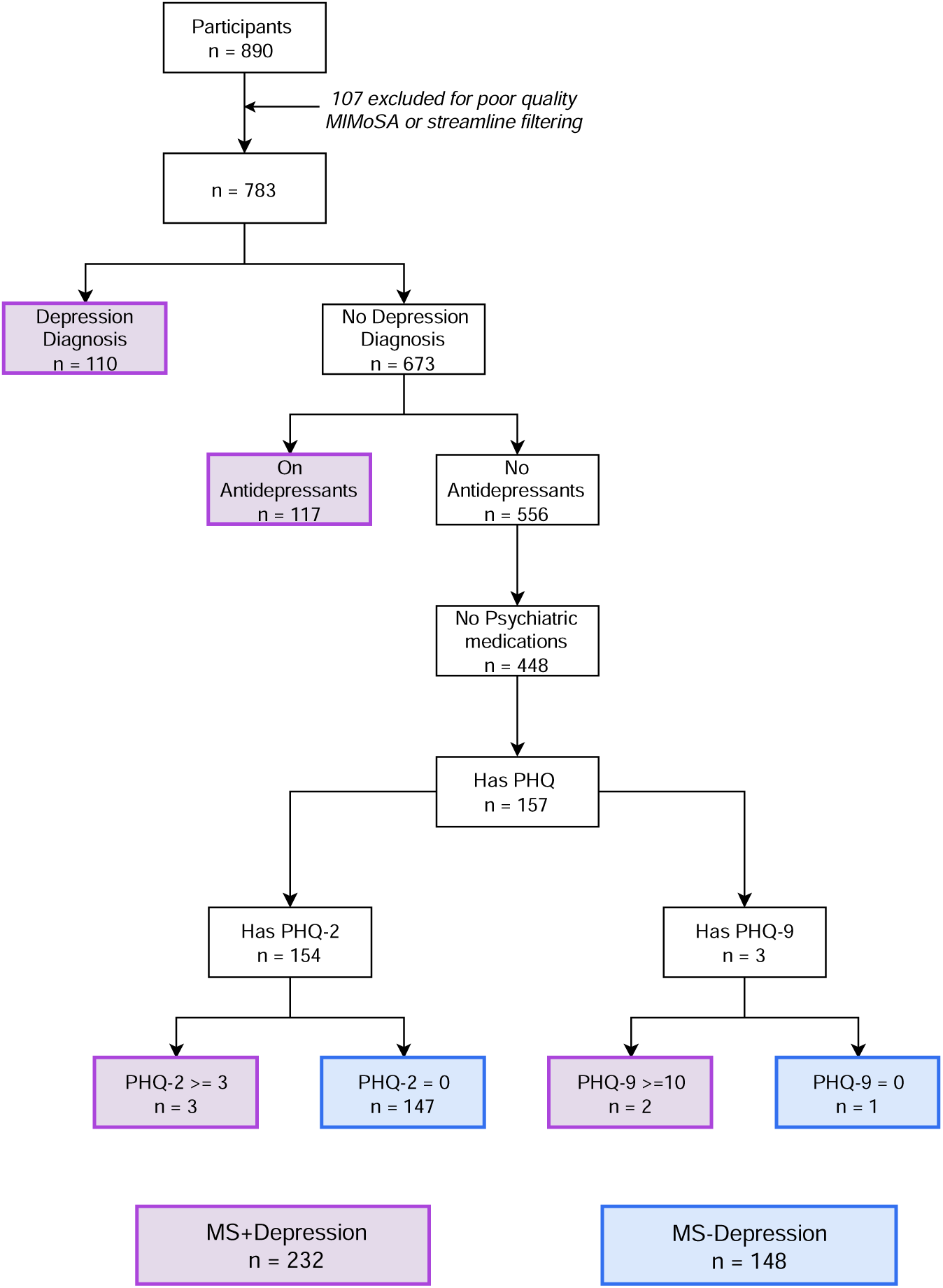
Flowchart of the study population. After excluding participants with poor quality scans, patients were stratified by depression diagnosis, prescription for psychiatric medications, and depression symptom screening. MIMoSA (Method for Inter-Modal Segmentation Analysis), PHQ (Patient Health Questionnaire), PHQ-2 (Patient Health Questionnaire-2), PHQ-9 (Patient Health Questionnaire-9), MS+Depression (Multiple sclerosis with depression), MS-Depression (Multiple sclerosis without depression).

#### Multiple sclerosis diagnosis

Participants were included if they received an ICD-10 MS diagnosis (G35) from a specialist in the University of Pennsylvania Comprehensive MS Center and received 3T MRI under the University of Pennsylvania MS protocol (detailed below). This sample was then stratified into samples of depressed (*MS+Depression*) and nondepressed comparators (*MS-Depression*).

#### Depression diagnosis

Given the heterogeneity in coding practices and the known underdiagnosis of depression in medical populations^28^, a multistep process was taken to classify our MS+Depression and MS-Depression samples. To be classified in the MS+Depression group, participants met one of three criteria: 1) ICD-10 code F32 (depressive episode), F33 (major depressive disorder), or F34 (persistent mood [affective] disorder); 2) screened positive for depression on PHQ-2 (>=3) or PHQ-9 (>=10); 3) previously prescribed an antidepressant medication^29^. Since the absence of an ICD-10 depression diagnosis does not exclude a diagnosis of depression, all MS-Depression patients had no previous depression diagnosis, were asymptomatic on PHQ-2 or 9 (score=0), and had no prescription history for psychiatric medications. Participants were excluded if they had an ICD-10 diagnosis of bipolar disorder or a manic episode, had no psychiatric or medication history indicating depression but never completed a screening PHQ-2 or PHQ-9, or were never formally evaluated by a Penn MS specialist.

To validate this classification, we evaluated group differences in the Patient Reported Outcomes Measurement Information System (PROMIS) scores in a subset of patients^30^. Notably, the PROMIS was not used in the definition of our groups, and thus was well-suited for group validation. The PROMIS assesses current symptom burden in 10 domains, including mental health and mood, emotional problems, quality of life, physical health, social activities satisfaction, carrying out social activities, carrying out physical activities, fatigue, overall physical impairment, and overall mental impairment. For patients who completed multiple PROMIS scales, the score most proximate to their imaging session was used.

### White matter depression network construction

To construct the white matter depression network, we identified fascicles that served as the structural backbone of a functional depression circuit map from Siddiqi et al^21^. Briefly, Siddiqi and colleagues evaluated correlations between depression and lesions or stimulation sites across 14 heterogeneous datasets and created an unthresholded mean correlation map. We first constructed a binary map by applying a threshold of *T*>3.09 to identify voxels with a statistically significant positive association between depression symptoms and brain disease or stimulation. We then constructed the white matter depression network using tools from DSI studio^31–33^. We built 77 canonical fascicles spanning cortical and subcortical regions from an atlas that was derived from a large sample of high-quality diffusion MRI (dMRI) and verified by neuroanatomists to correspond to known neuroanatomy^31^. We next calculated the volume (in voxels) that overlapped between the fascicle’s individual fibers, or streamlines, and the binarized functional depression network. Fascicles were ranked by their volume of overlap with the functional depression network. Fascicles with the highest degree of overlap (top 25%, 19 fascicles) were considered to be in the white matter depression network (**Figure 2; eTable 1**).

**Figure 2.**
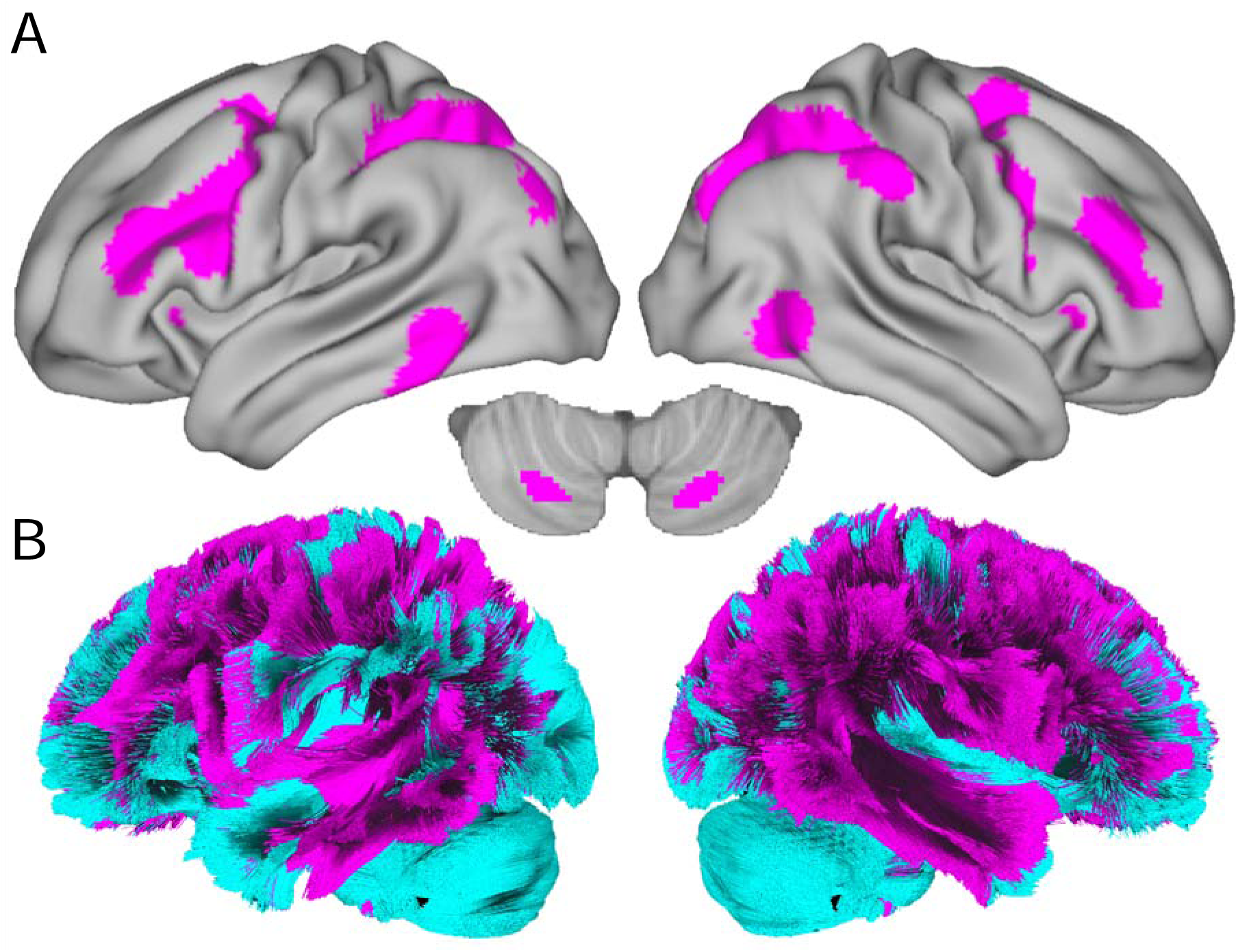
White matter depression network construction. A) The gray matter depression network from Siddiqi et al.^21^, thresholded at T>3.09. B) The white matter depression network was constructed from the top 25% of fascicles with the greatest volume of overlap with the gray matter depression network. Pink fascicles correspond to the depression network, whereas blue fascicles correspond to the nondepression network.

To ensure that our results were not driven by arbitrary distinctions between the depression and nondepression networks, we also constructed and tested white matter depression networks using alternate thresholds (e.g., the top 20% and 33%). We also repeated our primary analyses excluding the corpus callosum (CC) from the 25% depression network given the fascicle’s large size compared with other fascicles. Lastly, we summarized the lesion burden within each network (volume of impacted streamlines, divided by the total volume of streamlines within each network).

### Image acquisition and processing

#### Image acquisition & preprocessing

Structural MRI was obtained as part of routine care at 3T using a research-quality protocol, including 3D T1w MPRAGE (TR=1.9s, TE=2.48ms, TI=900ms, FA=9°, acquisition time=4:18, 176 sagittal slices, resolution=1mm^3^) and 3D T2 FLAIR (TR=5s, TE=398ms, TI=1.8s, FA=120°, acquisition time=5:02, 160 sagittal slices, resolution=1mm^3^). Images were processed using a previously described pipeline^34^. T1w and FLAIR images were N4-bias corrected^35^, extracerebral voxels were removed from the T1w images using Multi Atlas Skull Stripping (MASS)^36^, T1w images and their corresponding brain masks were registered to the corresponding FLAIR, and skull-stripped FLAIR and aligned T1w images were intensity normalized using WhiteStripe^37^. The quality of all processed images and segmentations were assessed by an imaging scientist with years of experience in MS imaging research.

### Automated Lesion Segmentation and Streamline Filtering

#### Automated lesion segmentation

Fully automated lesion segmentation was performed with the Method for Inter-Modal Segmentation Analysis (MIMoSA) to obtain binary maps of white matter lesions in a subject’s native space^38^. Prior work has demonstrated that MIMoSA performs similarly to manual segmentation^38^.

#### Analysis of white matter fascicles

To assess the impact of white matter lesions on fascicles across subjects, we performed streamline filtering in DSI Studio^31–33^. This required identifying whether individual streamlines within a fascicle were impacted (i.e., passed through a lesion), or spared (avoided a lesion). Delineating fascicles in a single dMRI dataset is known to be error-prone^39,34^, so we compared spatially-normalized lesions to canonical fascicles in template space.

Individual lesion maps (**eFigure 1A**) were transformed to the template space of the canonical fascicles (MNI2009bAsymmetric)^40^ using the T1w-based transform calculated by antsRegistration (**eFigure 1B**)^41,42^. Streamlines intersecting lesions at any point in their trajectory were considered injured and isolated from the rest of the fascicle. The total volume occupied by injured streamlines was calculated as the measure of disease burden in the fascicle (**eFigure 1C-E**). This was repeated for each of the 77 fascicles. For each individual, we also calculated the relative disease burden across all white matter as the volume of injured streamlines divided by the complete volume of all fascicles.

### Statistical Analyses

#### Primary analyses

Given the high comorbidity of depression in MS, we first assessed whether MS lesions were randomly distributed throughout the brain or preferentially targeted fascicles within the depression network, irrespective of diagnosis. Next, we explored whether MS+Depression patients had more lesion burden than MS-Depression patients. We then evaluated whether there was an interaction between network location and diagnosis. We tested these hypotheses with a linear mixed-effects model (R-package lme4 in R v3.2.5)^37^ that included fixed effects of network, diagnosis, network-by-diagnosis, and a random intercept for participants.

#### Secondary analyses

For our primary analyses, we defined the depression network as a binary map and assigned each fascicle to be either within or outside the depression network. However, it is also possible that the relationship is continuous, and disease in fascicles with greater overlap with the functional depression network are more likely to contribute to differences in depression diagnosis. To assess this, we evaluated disease within each fascicle separately. For each fascicle, we first calculated the volume (in voxels) of overlap of the fascicle with the functional depression network. Next, we computed the effect size (*r*) from a Wilcoxon signed-rank test comparing volume of disease in that fascicle between MS+Depression versus MS-Depression. Finally, we used a linear model to test for an association between each fascicle’s overlap with the functional depression network and the effect size from the MS+Depression versus the MS-Depression analysis.

#### Code availability

Code and instructions for replicating all analyses can be found at: https://pennlinc.github.io/msdepression/.

## RESULTS

Two hundred thirty-two MS patients with depression (MS+Depression; mean age[SD]=49[12], %females=86) and 148 age- and sex-matched nondepressed comparators (MS-Depression; age[SD]=47[13]; %females=79) were included in the analysis. As expected, MS+Depression patients had more depression symptoms than MS-Depression patients (**eTable 2**). A subsample of patients completed the PROMIS scales (MS+Depression n=49, MS-Depression n=36); this scale was not used to construct our sample and thus allowed us to independently validate our diagnostic groups. MS+Depression patients were more impaired than the MS-Depression sample across 9 of 10 PROMIS measures (**eTable 3**). However, there were no significant differences in fatigue between the diagnostic groups.

### Lesion burden is higher in both the depression network and in MS+Depression patients

Across all patients with MS, lesion burden preferentially affected fascicles within versus outside the white matter depression network (β=0.09, 95% CI=0.08-0.10, P<0.001; **Figure 3**). A main effect of diagnosis was also noted, where MS+Depression patients had more lesion burden across the whole brain as compared to MS-Depression patients (β=0.06, 95% CI=0.01-0.10, P=0.015; **Figure 4A**). We next tested whether the diagnostic differences between MS+Depression and MS-Depression patients were network-specific. We found a network-by-diagnosis interaction (β=0.02, 95% CI=0.003-0.040, P=0.020; **Figure 4B**), which was specifically driven by worse lesion burden in MS+Depression within the depression network. In addition to comparing disease burden between MS+Depression and MS-Depression at the network level, we compared fascicle-level burden between diagnostic groups (**Figure 5A**). We found that fascicles with more overlap with the depression network also had greater disease burden when comparing MS+Depression to MS-Depression patients (**Figure 5B**; Adjusted *R*^2^=0.06, P_lm_=0.02).

**Figure 3.**
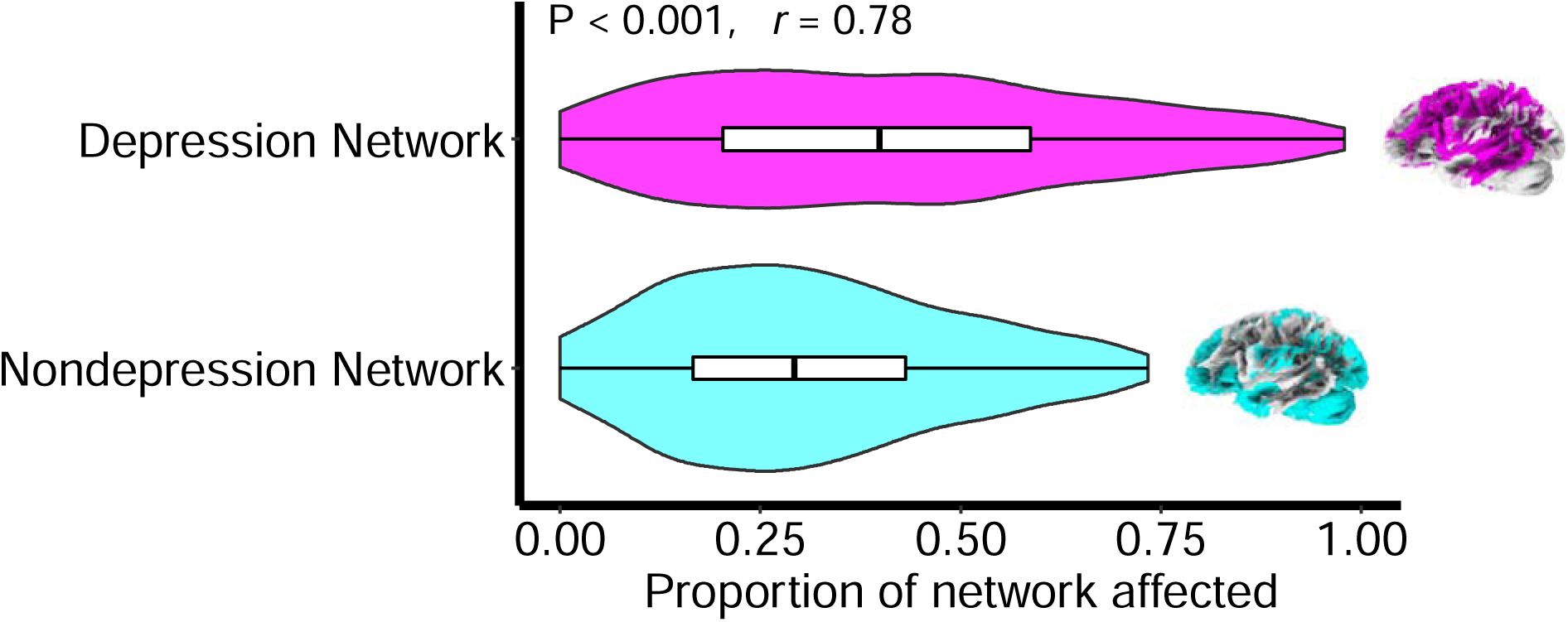
Multiple sclerosis lesions preferentially impacted white matter fascicles in the depression network. Patients with MS have enrichment of disease in fascicles that connect areas in the functional depression network (P<0.001, *r*=0.78).

**Figure 4.**
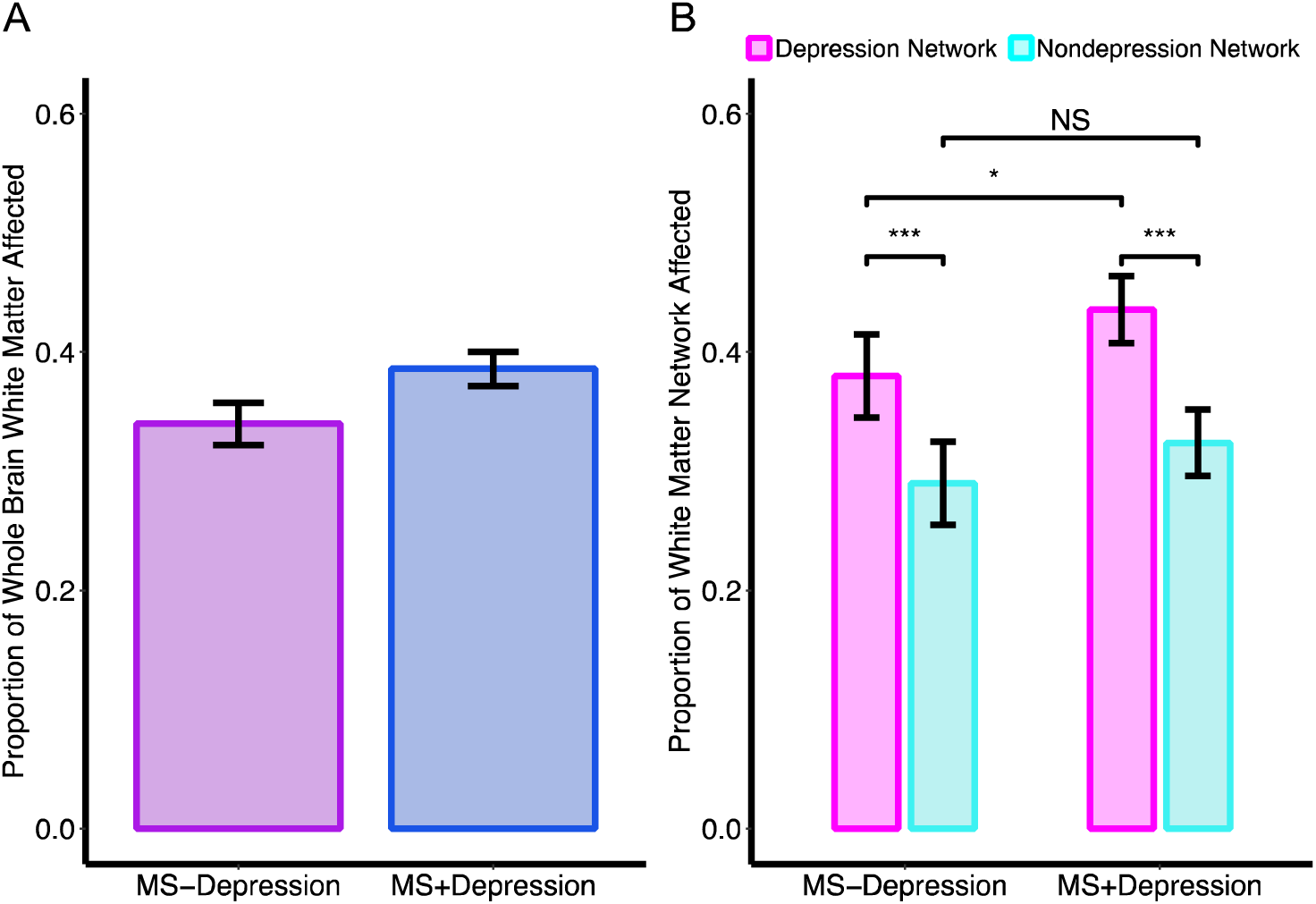
Multiple sclerosis patients with depression (MS+Depression) have more disease than multiple sclerosis patients without depression (MS-Depression) in the depression network. A) Across the whole brain, MS+Depression patients have more white matter disease burden than MS-Depression patients (P=0.04). B) A network-by-diagnosis interaction was noted (P=0.02), which is driven by worse disease in MS+Depression patients, specifically in the depression network. * P<0.05, *** P<0.001, NS=Nonsignificant.

**Figure 5.**
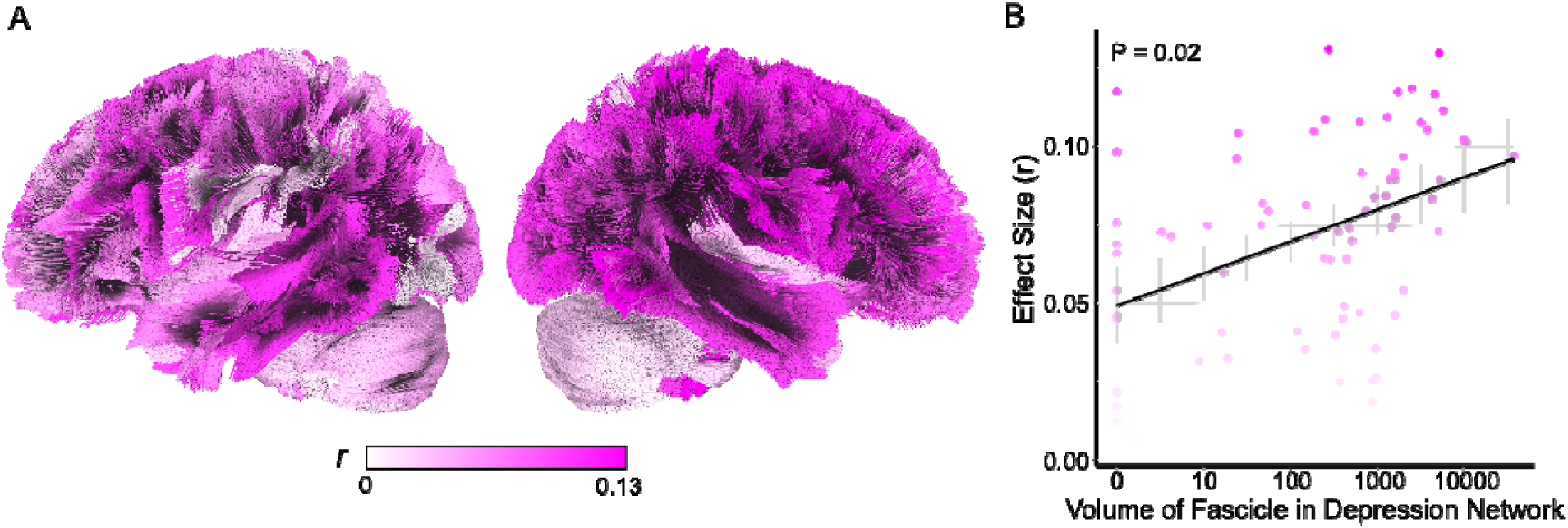
Between diagnosis differences in disease burden are larger in fascicles with greater overlap with the depression network. A) Individual fascicles are colored by the effect size from a Wilcoxon signed-rank test comparing disease burden in MS+Depression versus MS-Depression patients, with darker colors indicating more severe disease in MS+Depression patients. B) Larger effect sizes in the between-diagnosis Wilcoxon signed-rank test are associated with greater overlap of the fascicle with the depression network (P=0.02). The x-axis is logarithmically scaled for visualization purposes.

### Convergence of results across alternative methods for defining white matter depression network

To ensure that our results were not driven by the threshold we used to define fascicles within the depression network (e.g., top 25%), we repeated our analyses using alternate thresholds for assigning fascicles to the depression network (top 33% or top 20%). We also repeated our analysis excluding the corpus collosum given its large size compared with other fascicles. In all analyses, our results remained significant. Using a 33% threshold, we found a main effect of network (β=0.122, 95% CI=0.107-0.138, P<0.001) and network-by-diagnosis interaction (β=0.022, 95% CI=0.002-0.042, P=0.029). Similarly, with a 20% threshold, we still found a main effect of network (0.110, 95% CI=0.10-0.12, P<0.001) and network-by-diagnosis interaction (β=0.022, 95% CI=0.004-0.040, P=0.018). Convergence was also seen in analyses that excluded the corpus collosum: main effect of network (β=0.03, 95% CI=0.02-0.05, P<0.001) and network-by-diagnosis interaction (β=0.02, 95% CI=0.002-0.040, P=0.03). Taken together, we demonstrate that our results are robust across multiple depression network definitions.

## DISCUSSION

Using a novel approach for assessing the relationship between white matter lesions and brain networks implicated in depression, we provide new evidence supporting an association between white matter lesion location and depression in MS. Irrespective of depression diagnosis, patients with MS had greater disease burden within the white matter depression network. This anatomical predilection may create a vulnerability to depression comorbidity in MS; patients with MS+Depression had higher disease burden across the whole brain than MS-Depression patients and greater burden specifically within the white matter depression network. Additionally, MS patients with depression had greater injury in fascicles with more overlap with the depression network. Taken together, these data demonstrate that injury to white matter fascicles that structurally support a previously defined functional depression network^21^ are associated with depression in MS.

Until recently, numerous conceptual and methodological challenges have limited the field’s understanding of how MS lesions may increase vulnerability to depression. Prior studies built on the assumption that depression symptoms are caused by lesions in the same anatomic location have yielded inconsistent findings, indicating that spatially distributed lesions can lead to a common phenotype^8,17,43^. However, few studies have directly explored whether heterogeneous lesions of the same network, rather than the same location, contribute to depression. Large sample sizes are necessary to identify complex relationships between distributed white matter lesions and depression, but previous studies often employed manual lesion segmentation to extract white matter lesions from brain scans, which is time-intensive and subject to bias^39^. Given the high resolution necessary for segmentation, prior work often relied on research scans, increasing costs and effort for recruitment^43,44^. Together, these constraints have limited efforts to disentangle the complex relationship between MS and depression.

To address these limitations, our analysis coupled automated lesion segmentation in clinical scans with white matter lesion network mapping to show that lesions in white matter fascicles connecting the depression network are associated with depression in MS patients. These fascicles connect a previously described functional depression network that includes the frontoparietal and dorsal attention networks, which support executive function and attention. Importantly, planning and concentration difficulties are core deficits in depression^45–48^. Furthermore, the dorsolateral prefrontal cortex, a central brain region in the frontoparietal network, is a structural target for transcranial magnetic stimulation and electroconvulsive therapy for severe depression^49,50^. Disease in these fascicles may therefore impact treatment outcomes and warrant further study.

In recent work, Siddiqi et al. used normative data to estimate the relationship between BOLD signal in MS white matter lesions to the functional depression network^22^. They showed associations between functional connectivity and variation in depressive symptoms in a sample centered on nondepressed individuals. We extend this literature by using white matter lesions to define participant-specific, disconnected white matter networks and directly test whether structural injury to key fascicles relates to a history of clinically-impairing depression. Additionally, we capitalized on a new dataset of research-quality clinical scans to increase sample size and generalizability. Lastly, our conceptual framework complements recent literature showing that MS-associated structural disruptions are associated with disability and disease progression^23,24,51,52^.

### Limitations

Our study uses ICD-10 diagnostic coding for both depression and MS. For depression phenotyping, ICD-10 diagnosis reflects a lifetime history of depression rather than symptoms at the time of scan. Depression is underdiagnosed in medical populations––the absence of an ICD-10 depression code does not preclude the presence of depression^28^. Our rigorous, multistep depression phenotyping that incorporated medications and symptom screenings aimed to address this limitation, and was validated by an independent measure not used in group construction (i.e., PROMIS). Future prospective studies with structured depression assessments will likely yield additional insights into the relationship between active depressive symptoms and white matter lesion location and burden. Additionally, though this study characterizes MS as a disease of white matter, MS is also associated with gray matter atrophy^53–55^. Incorporating measures of gray matter disease into future studies of MS and depression will likely be informative. Lastly, medications for multiple sclerosis (i.e., steroids) have been associated with psychiatric symptoms^56^. Prospective studies that relate the initiation of MS treatments with depression are warranted.

### Generalizability

We demonstrate that lesions to white matter fascicles can contribute to depression. Our work highlights opportunities to combine clinical imaging and EHR data to capture individual variation related to depression. By using automated white matter lesion segmentation, we have provided a scalable solution for expanding this work to bigger datasets, as well as datasets outside our hospital system. Additionally, our template-based approach does not require diffusion-weighted MRI, allowing for wider use.

### Conclusions

In this retrospective case-control study, we sought to explore the high comorbidity between MS and depression using white matter lesion network mapping. We identified key relationships between white matter lesions in the depression network and depression. MS lesions preferentially target white matter fascicles that connect the depression network irrespective of depression diagnosis. Furthermore, MS patients with depression have more disease burden than nondepressed MS patients, which is specifically driven by more disease burden within the white matter depression network. This approach holds promise not just for understanding depression in the context of MS, but the role of abnormalities in white matter as a mechanism for depression more broadly.

## Data Availability

All data produced in the present work are contained in the manuscript. Though raw files cannot be released due to HIPAA protections, all code have been made public on github.

https://pennlinc.github.io/msdepression/

## Author Contributions

Drs. Baller, Shinohara, and Satterthwaite had full access to all the data in the study and take responsibility for the integrity of the data and the accuracy of the data analysis.

### Concept and design

Baller, Sweeney, Cieslak, Schindler, Shinohara, Satterthwaite

### Acquisition, analysis, or interpretation of data

Baller, Sweeney, Cieslak, Covitz, Martin, Schindler, Bar-Or, Markowitz, Manning, Perrone, Rautman, Detre, Fox, Shinohara, Satterthwaite.

### Drafting of the manuscript

Baller, Satterthwaite.

### Critical revision of the manuscript for important intellectual content

Baller, Sweeney, Cieslak, Schindler, Shinohara, Satterthwaite.

### Statistical analysis

Baller, Sweeney.

### Obtained funding

Baller, Shinohara, Satterthwaite.

### Administrative, technical, or material support

Elahi, Rautman, Seitz

### Supervision

Shinohara, Satterthwaite

## Data sharing

The clinical dataset taken from the University of Pennsylvania is strictly private and cannot be shared.

## Online-only Supplement

### Supplemental Material

**eTable 1.** Fascicles ranked by volume of overlap with the functional depression network. The white matter depression network was composed of the top 25% (19 fascicles) with the greatest overlap with the functional depression network (highlighted in gray).

| Fascicle | Volume (number of voxels) in functional depression mask |
| --- | --- |
| Corpus Callosum | 37726 |
| Arcuate Fasciculus (Left) | 10677 |
| Superior Longitudinal Fasciculus 2 (Right) | 10064 |
| Superior Longitudinal Fasciculus 3 (Right) | 5780 |
| Frontal Aslant Tract (Left) | 5246 |
| Thalamic Radiation Superior (Left) | 5142 |
| Superior Longitudinal Fasciculus 2 (Left) | 5050 |
| Arcuate Fasciculus (Right) | 4616 |
| Thalamic Radiation Superior (Left) | 4252 |
| Frontal Aslant Tract (Right) | 3759 |
| Corticostriatal Tract Superior (Right) | 3167 |
| Corticopontine Tract Frontal (Right) | 2485 |
| Corticopontine Tract Parietal (Left) | 1993 |
| Inferior Longitudinal Fasciculus (Right) | 1978 |
| Corticopontine Tract Parietal (Right) | 1727 |
| Corticostriatal Tract Superior (Left) | 1643 |
| Parietal Aslant Tract (Right) | 1607 |
| Middle Longitudinal Fasciculus (Left) | 1598 |
| Corticobulbar Tract (Right) | 1576 |
| Corticostriatal Tract Posterior (Right) | 1468 |
| Parietal Aslant Tract (Left) | 1383 |
| Cortico Spinal Tract (Right) | 1299 |
| Inferior Fronto Occipital Fasciculus (Right) | 1261 |
| Cingulum Parahippocampal Parietal (Right) | 980 |
| Middle Longitudinal Fasciculus (Right) | 964 |
| Corticopontine Tract Occipital (Right) | 934 |
| Superior Longitudinal Fasciculus 3 (Left) | 886 |
| Vermis | 871 |
| Corticobulbar Tract (Left) | 746 |
| Thalamic Radiation Posterior (Left) | 658 |
| Thalamic Radiation Posterior (Right) | 625 |
| Extreme Capsule (Right) | 619 |
| Corticostriatal Tract Posterior (Left) | 512 |
| Corticopontine Tract Frontal (Left) | 476 |
| Inferior Fronto Occipital Fasciculus (Left) | 445 |
| Thalamic Radiation Anterior (Left) | 417 |
| Corticostriatal Tract Anterior (Left) | 396 |
| Cerebellum (Right) | 373 |
| Reticulospinal Tract (Right) | 357 |
| Extreme Capsule (Left) | 329 |
| Corticostriatal Tract Anterior (Right) | 293 |
| Dentatorubrothalamic Tract (Right) | 277 |
| Thalamic Radiation Anterior (Right) | 250 |
| Cortico Spinal Tract (Left) | 245 |
| Corticopontine Tract Occipital (Left) | 186 |
| Superior Longitudinal Fasciculus 1 (Right) | 183 |
| Superior Longitudinal Fasciculus 1 (Left) | 152 |
| Medial Lemniscus (Left) | 148 |
| Reticulospinal Tract (Left) | 120 |
| Acoustic Radiation (Right) | 56 |
| Dentatorubrothalamic Tract (Left) | 48 |
| Cingulum Parahippocampal Parietal (Left) | 46 |
| Vertical Occipital Fasciculus (Right) | 25 |
| Anterior Commissure | 24 |
| Middle Cerebellar Peduncle | 19 |
| Cingulum Frontal Parietal (Left) | 17 |
| Cerebellum (Left) | 16 |
| Acoustic Radiation (Left) | 11 |
| Cingulum Frontal Parietal (Left) | 9 |
| Inferior Longitudinal Fasciculus (Left) | 4 |
| Fornix (Right) | 3 |
| Cingulum Parolfactory (Right) | 1 |
| Cingulum Frontal Parahippocampal (Left) | 0 |
| Cingulum Frontal Parahippocampal (Right) | 0 |
| Cingulum Parahippocampal (Left) | 0 |
| Cingulum Parahippocampal (Right) | 0 |
| Cingulum Parolfactory (Left) | 0 |
| Uncinate Fasciculus (Left) | 0 |
| Uncinate Fasciculus (Right) | 0 |
| Vertical Occipital Fasciculus (Left) | 0 |
| Inferior Cerebellar Peduncle (Left) | 0 |
| Inferior Cerebellar Peduncle (Right) | 0 |
| Superior Cerebellar Peduncle | 0 |
| Fornix (Left) | 0 |
| Medial Lemniscus (Right) | 0 |
| Optic Radiation (Left) | 0 |
| Optic Radiation (Right) | 0 |

**eTable 2.** Participant Demographics. Sex and race/ethnicity variables were extracted from the electronic health record. Race and ethnicity, both coded under the field “Race,” are patient-reported. P values reflect T-tests in continuous data (Age) and chi square for categorical variables (Sex, Race/Ethnicity).

|  | MS+Depression | MS-Depression | P |
| --- | --- | --- | --- |
| n | 232 | 148 |  |
| Age (mean [SD]) | 48.8 [11.8] | 46.6 [12.7] | 0.09 (NS) |
| Sex (% Female) | 199 (85.8%) | 117 (79.1%) | 0.12 (NS) |
| Race and Ethnicity |  |  | 0.60 (NS) |
| American Indian | 0 (0%) | 2 (1.4%) |  |
| Asian | 2 (0.9%) | 1 (0.7 %) |  |
| Black | 42 (18.1%) | 28 (18.9%) |  |
| East Indian | 0 (0%) | 1 (0.7%) |  |
| Hispanic/Latino/Black | 1 (0.4%) | 0 (0%) |  |
| Hispanic/Latino/White | 3 (1.3%) | 3 (2.0%) |  |
| Other | 8 (3.4%) | 3 (2.0%) |  |
| Unknown | 3 (1.3%) | 2 (1.4%) |  |
| White | 173 (74.6%) | 108 (73.0%) |  |

**eTable 3.** Reported Outcomes Measurement Information System (PROMIS) scores. Lower scores represent more impairment. SD = standard deviation. P values reflect T-tests in continuous data (Age, PROMIS scores) and chi-squared for categorical variables (Sex, Race).

|  | MS+Depression | MS-Depression | P |
| --- | --- | --- | --- |
| n | 49 | 36 |  |
| Age (mean [SD]) | 50.0 [10.9] | 54.0 [11.4] | 0.11 (NS) |
| Sex (% Female) | 41 (83.7%) | 31 (86.1%) | 1.0 (NS) |
| Race (% White) | 32 (65.3%) | 26 (72.2%) | 0.66 (NS) |
| Quality of Life (mean [SD]) | 3.0 [1.0] | 3.6 [1.0] | 0.010 |
| Physical Health (mean [SD]) | 2.4 [0.8] | 2.9 [0.8] | 0.019 |
| Mental Health and Mood (mean [SD]) | 3.4 [1.0] | 4.0 [0.8] | 0.011 |
| Social Activities Satisfaction (mean [SD]) | 2.9 [1.4] | 3.8 [1.0] | 0.001 |
| Carrying out Social Activities (mean [SD]) | 2.9 [1.2] | 3.7 [1.1] | 0.007 |
| Carrying out Physical Activities (mean [SD]) | 3.1 [1.0] | 3.7 [1.1] | 0.032 |
| Emotional Problems (mean [SD]) | 3.0 [1.1] | 3.7 [1.0] | 0.011 |
| Fatigue Average (mean [SD]) | 3.1 [1.1] | 3.5 [1.3] | 0.18 (NS) |
| Postoperative Physical Score (mean [SD]) | 11.7 [2.6] | 13.2 [3.0] | 0.032 |
| Postoperative Mental Score (mean [SD]) | 12.3 [3.3] | 15.0 [2.9] | 0.001 |

**eFigure 1.**
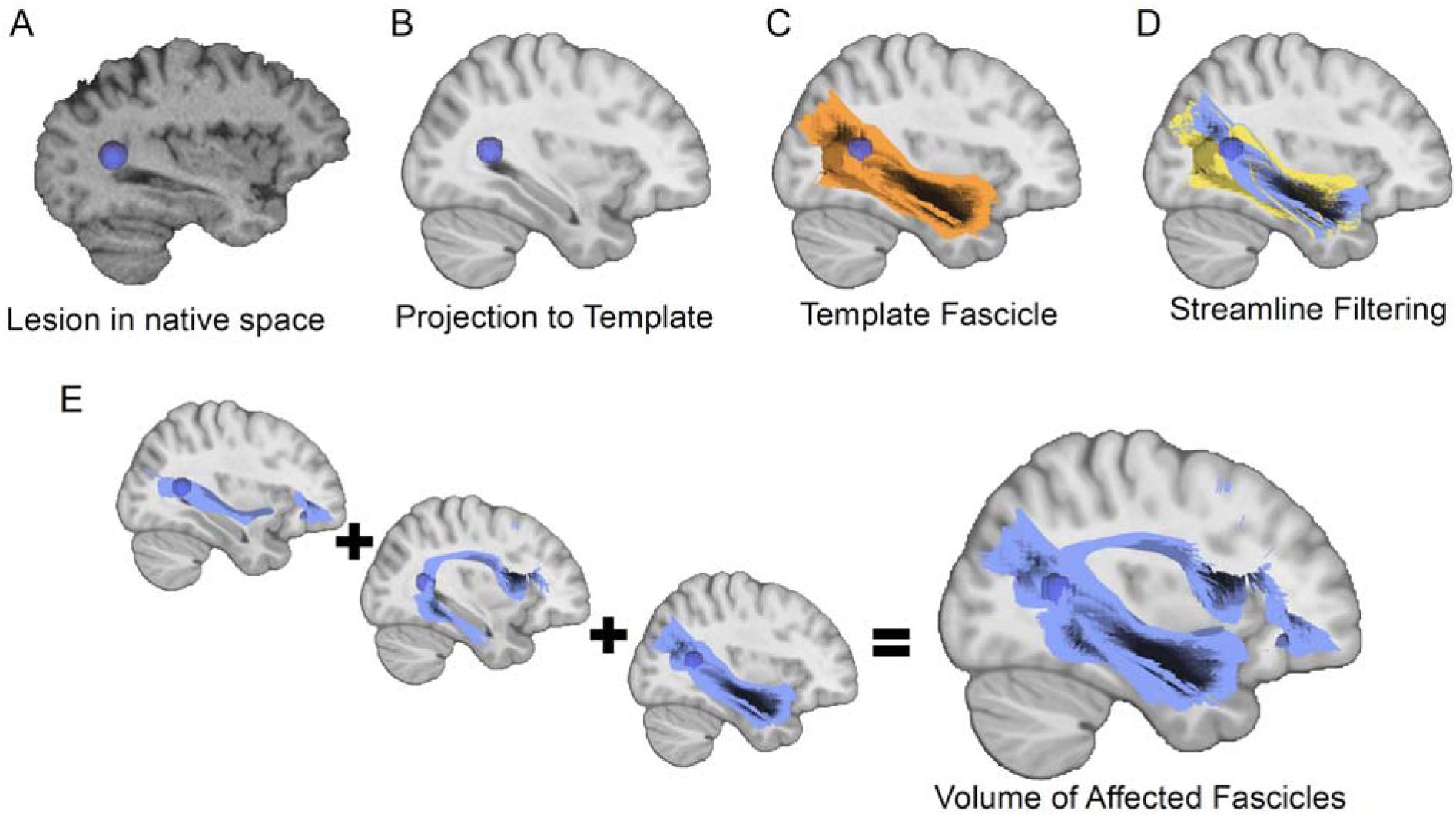
Fascicle analysis pipeline. A) Lesions were segmented with MIMoSA and B) transformed to the template space of the canonical fascicles (MNI2009bAsymmetric)^40^. C-D) For each fascicle, streamlines intersecting lesions at any point in their trajectory were considered injured and isolated from the rest of the fascicle (blue). The total volume occupied by injured streamlines was calculated as the measure of disease burden in the fascicle. E) This was repeated at each of 77 fascicles to obtain measures of disease burden.

